# Combinatorial language parent-report score differs significantly between typically developing children and those with Autism Spectrum Disorders

**DOI:** 10.1101/2022.03.30.22271503

**Authors:** Matthew Arnold, Andrey Vyshedskiy

**Author notes:** Corresponding author: Andrey Vyshedskiy, PhD, Boston University, Boston, MA, USA.

## Abstract

Prefrontal synthesis (PFS) is a component of constructive imagination. It is defined as the process of mentally juxtaposing objects into novel combinations. For example, to comprehend the instruction “put the cat under the dog and above the monkey,” it is necessary to use PFS in order to correctly determine the spatial arrangement of the cat, dog, and monkey with relation to one another. The acquisition of PFS hinges on the use of combinatorial language during early development in childhood. Accordingly, children with developmental delays exhibit a deficit in PFS, and frequent assessments are recommended for these individuals. In 2018, we developed the Mental Synthesis Evaluation Checklist (MSEC), a parent-reported evaluation designed to assess PFS and combinatorial language comprehension. In this manuscript we use MSEC to identify differences in combinatorial language acquisition between ASD (N=29138) and neurotypical (N=111) children. Results confirm the utility of the MSEC in distinguishing language deficits in ASD from typical development as early as 2 years of age (p<0.0001).

## Introduction

When a receptive language test measures the number of words children know, it primarily evaluates the ability of the Wernicke’s area to match words to mental images (Friederici, 2011). Conversely, to assess combinatorial receptive language, one must evaluate the ability of the lateral prefrontal cortex (LPFC) to synthesize novel mental images from objects stored in memory in accordance with imposed grammatical rules (Vyshedskiy, 2019). It is this function of the LPFS that we have named prefrontal synthesis (PFS), formerly known as mental synthesis (Vyshedskiy, 2008; Vyshedskiy & Dunn, 2015). PFS is neurologically different from other key components of mental imagery, such as dreaming and simple memory recall. Unlike dreaming, which is not controlled by the LPFC and occurs spontaneously (Braun et al., 1997; Solms, 1997), PFS is completely dependent on an intact LPFC (Baker et al., 1996; Christoff & Gabrieli, 2000; Duncan et al., 1995; Fuster, 2008; Luria, 2012; Waltz et al., 1999). Furthermore, simple memory recall differs from PFS as well, as it involves the recall of a single object encoded at some point in the past, whereas PFS involves the recollection and novel combination of two or more objects encoded in one’s memory (Vyshedskiy, Dunn, et al., 2017; Vyshedskiy & Dunn, 2015).

PFS is defined narrowly in order to separate it from other components of executive function, such as working memory, impulse control, attention, and cognition (Vyshedskiy, Khokhlovich, et al., 2020). An important distinction must be made that PFS is not synonymous with problem-solving. This is because, complex problems can often be solved via amodal completion (Gerbino & Salmaso, 1987; Weigelt et al., 2007), spontaneous insight (Salvi et al., 2016), integration of modifiers (Vyshedskiy, Dunn, et al., 2017), and various other mechanisms that either do not require the LPFC and/or do not involve the voluntary combining of objects stored in one’s memory (Pearson, 2019).

PFS is acquired by neurotypical individuals around 4 years of age (Vyshedskiy, DuBois, et al., 2020), but acquisition of PFS is known to be a common challenge for children with intellectual disabilities. PFS paralysis has been previously categorized as “stimulus overselectivity,” “tunnel vision,” and “lack of responsivity to multiple clues” (Lovaas et al., 1979; Ploog, 2010; Schreibman, 1988).

Impaired PFS reduces understanding of combinatorial language and complex arithmetic, and can affect virtually every aspect of an individual’s verbal, cognitive, and social functioning (Vyshedskiy, Mahapatra, et al., 2017). Furthermore, independent life is nearly impossible without understanding of spatial prepositions, recursion, and complex arithmetic (Bigby, 1996). Thus, combinatorial language is commonly the ultimate concern of parents, as they want their children to be able to live an independent life (Finke et al., 2019).

Language therapists are cognizant of the short critical period for PFS acquisition, and there is a wide consensus that intense early intervention should be administered to children as soon as possible to help alleviate speech impairments early on (Dawson et al., 2010). The goals of speech language pathologists and applied behavior analysis therapists also happen to be built around the construct of PFS. As a result, these professionals are taught to focus their efforts on the treatment of impaired PFS. Some examples of PFS developing techniques as described by speech language pathologists are “combining adjectives, location/orientation, color, and size with nouns,” “following directions with increasing complexity,” and “building the multiple features/clauses in the sentence” (American Speech-Language-Hearing Association, 2016). In applied behavior analysis jargon, these techniques are known as “visual-visual and auditory-visual conditional discrimination” (Axe, 2008; Eikeseth & Smith, 2013; Lowenkron, 2006; Michael et al., 2011), “development of multi-cue responsivity” (Lovaas et al., 1979), and “reduction of stimulus overselectivity” (Ploog, 2010).

Despite widespread recognition for the importance of early development of combinatorial language, there is a clear lack of psychometric tests that are able to measure this ability in early language learners at the level of 3 to 4-year-old children. Existing tests for combinatorial language – the Preschool Language Scales (PLS-5) (Zimmerman et al., 2011), the Token Test for children (A. De Renzi & Vignolo, 1962; E. De Renzi & Faglioni, 1978), and the Test for Reception Of Grammar (TROG) (Bishop, 2005) – are geared for children age five and older. For example, the easiest question assessing spatial prepositions in PLS-5 (Q39: “Put the Mr. Bear in back of you,” “Put the Mr. Bear in front of me”) uses the combination of spatial prepositions with pronouns and is targeted to 4-and-a-half-year-old children. The next combinatorial language comprehension question after Q39 is Q46, that tests understanding of nested sentences and targets 6-year-old children.

Similarly, the Token Test uses overly long instructions that makes it too complex for early language learners. The simplest Token Test spatial preposition task instructions are: “Put the red circle on the green square” and “Put the green triangle on the red circle” (A. De Renzi & Vignolo, 1962; E. De Renzi & Faglioni, 1978). This instruction is twice as long as necessary to test comprehension of spatial prepositions (the simplest possible spatial preposition instruction only includes a subject, object, and the spatial preposition: e.g., “Put the lion on the giraffe”). Long instructions of the Token Test make the test inappropriate for young language learners and ASD children who commonly have short auditory memory (Edmunds et al., 2021).

The test for Reception of Grammar (TROG) has a variety of combinatorial language questions, but requires participants to answer by pointing to a correct picture, rather than showing the answer with tangible toy objects. Children with ASD and attention deficit disorder often fail to interpret picture-answers (Gomez et al., 2018). As a result, none of the existing combinatorial language tests are suited for assessing early language learners, particularly children with ASD and attention deficit disorder.

Accordingly, we have developed two combinatorial receptive language tests targeted at the level of 3- to 4-year-old children. The first is the clinician-administered 10-item Linguistic Evaluation of Prefrontal Synthesis (LEPS) that uses toys and the simplest and shortest combinatorial language instructions to assess PFS, such as; “Put the giraffe under the monkey,” “Put the elephant on top of the giraffe” (Vyshedskiy, DuBois, et al., 2020). Answering these questions involves arranging tangible objects and therefore avoids the problem with picture-answers interpretation (Gomez et al., 2018). The second test is the parent-reported 20-item Mental Synthesis Evaluation Checklist (MSEC) (Braverman et al., 2018). The MSEC uses questions about children’s behavior and sentence comprehension to elucidate their understanding of combinatorial language. The MSEC was previously validated with 3715 parents of children with ASD and demonstrated excellent psychometric qualities: good internal reliability, adequate test–retest reliability, construct validity, and known group validity (Braverman et al., 2018). In this manuscript we use MSEC to characterize combinatorial language and PFS acquisition in normally developing and ASD children 2 to 7 years of age. We were particularly interested in how early MSEC was able to identify the difference between normally developing and ASD children.

## Methods

### Neurotypical participants

A convenience sample of 111 neurotypical participants was obtained for this study by approaching parents of young children on a parent community online site and asking if they would be willing to complete a Google form. The data presented in this manuscript includes everyone who agreed to participate and indicated that their child was “Normally Developing” (other diagnostic options included: Mild Language Delay, Attention Deficit Disorder, Autism Spectrum Disorder, Asperger Syndrome, Social Communication Disorder, Specific Language Impairment, Apraxia, Sensory Processing Disorder, Down Syndrome, and Other). All participants’ caregivers had at least college education, 44% at least master’s, and 8% doctorate. All caregivers consented to anonymized data analysis and publication of the results. The mean age of participants was 4.8±1.7 (range, 2–8) years, and 48% of them were male. The breakdown of sample size within each age bin is indicated in Table 1.

**Table 1.** The breakdown of sample size within each age bin.

| **Age** | **Neurotypical** | **ASD** |
| --- | --- | --- |
| 2-2.9 | 17 | 5318 |
| 3-3.9 | 21 | 14104 |
| 4-4.9 | 13 | 15386 |
| 5-5.9 | 22 | 11763 |
| 6-6.9 | 22 | 8103 |
| 7-7.9 | 16 | 5512 |
| **Total** | **111** | **60186** |

### Participants with ASD

The MSEC was administered to users of a language therapy app. The app was made available *gratis* at all major app stores in September 2015. Once the app was downloaded, parents were asked to register and to provide demographic details, including the child’s diagnosis and age. Caregivers consented to anonymized data analysis and completed the Mental Synthesis Evaluation Checklist (MSEC), as well as several other assessments. Here we analyzed data from 29138 app users who identified their child’s diagnosis as ASD and completed 60186 evaluations (with an average of 2.1 evaluations per user). Other diagnostic options included: Mild Language Delay, Pervasive Developmental Disorder, Attention Deficit Disorder, Asperger Syndrome, Social Communication Disorder, Specific Language Impairment, Apraxia, Sensory Processing Disorder, Down Syndrome, Lost Diagnosis of Autism or PDD, Other Genetic Disorder, normally developed child. ASD condition was identified by 88% of responders. All participants who identified their diagnosis as ASD were included in the study. The education level of the participants’ caregivers’ was as follows: 94% had at least high school, 73% had at least college education, 35% at least master’s, and 5% doctorate. All caregivers consented to anonymized data analysis and publication of the results. The mean age of participants was 4.8±1.4 (range, 2–8) years, and 79% of them were male. The breakdown of sample size within each age bin is indicated in Table 1.

### Mental Synthesis Evaluation Checklist

The Mental Synthesis Evaluation Checklist (MSEC) test was introduced in 2018 to provide parents the ability to assess combinatorial receptive language and PFS (Braverman et al., 2018). The MSEC is rooted in a set of common language comprehension items. The purpose of each item is to determine whether or not an individual could mentally modify a single object and to combine several mental objects together, thereby indicating the level of overall PFS ability. Table 2 describes 20 MSEC items. Each item is assigned a score from 0 to 2 based on three possible answer choices: not true=2, somewhat true=1, and very true=0. Since the MSEC was developed to be complementary to the Autism Treatment Evaluation Checklist (ATEC) (Rimland & Edelson, 1999), it uses the same answer choices and the scoring system wherein a lower score indicates improvement. The total score is the sum of all items, thus, the maximum MSEC score is 40 and the minimum score is 0.

**Table 2.** MSEC items sorted by domains. For each item, a caregiver is asked how much each of the following is true regarding his/her child on a scale: not true (2 points), somewhat true (1 point), and very true (0 points).

| My child: | Domain of PFS |
| --- | --- |
| Understands some simple modifiers (i.e., green apple vs. red apple or big apple vs. small apple) | Sentence structure: understanding of noun-adjective combinations, spatial prepositions, and flexible syntax |
| Understands several modifiers in a sentence (i.e., small green apple) |  |
| Understands size (can select the largest/smallest object out of a collection of objects) |  |
| Understands possessive pronouns (i.e., your apple vs. her apple) |  |
| Understands spatial prepositions (i.e., put the apple ON TOP of the box vs. INSIDE the box vs. BEHIND the box) |  |
| Understands verb tenses (i.e., I will eat an apple vs. I ate an apple) |  |
| Understands the change in meaning when the order of words is changed (i.e., understands the difference between 'a cat ate a mouse' vs. 'a mouse ate a cat') |  |
| Understands simple stories that are read aloud | Narrative comprehension: understanding of stories and explanations |
| Understands elaborate fairy tales that are read aloud (i.e. stories describing FANTASY creatures) |  |
| Understands explanations about people, objects or situations beyond the immediate surroundings (e.g., "Mom is walking the dog," "The snow has turned to water") |  |
| Draws a VARIETY of RECOGNIZABLE images (objects, people, animals, etc.) | Creative manifestations: drawing and make-believe activities |
| Can draw a NOVEL image following YOUR description (e.g. a three-headed animal) |  |
| Engages in a VARIETY of make-believe activities (such as playing house, playing with toy soldiers, building forts and castles, etc.) |  |
| Understands NUMBERS (i.e. two apples vs. three apples) | Simple arithmetic: number manipulations with increasing complexity |
| Can perform simple arithmetic: $2 + 3 = ?$ | |
| Can add larger numbers: $7 + 6 = ?$ | |
| Can perform simple subtraction: $3 - 2 = ?$ | |
| Can subtract larger numbers: $15 - 7 = ?$ | |
| Can perform simple multiplication: $2 \times 2 = ?$ | |
| Can multiply larger numbers: $6 \times 7 = ?$ | |

## Results

Parents completed MSEC assessments for 29138 ASD children and 111 neurotypical children. Cross-sectional MSEC scores are shown in Figure 1. In neurotypical children MSEC score decreases near linearly, from 22.8±4.2 points at 2 (range 2 to 2.9) years of age to 2.7±2.5 points at 7 (range 7 to 7.9) years of age, with a lower score indicating improvement. Conversely, in ASD children the MSEC score starts at 32.5±7.9 points at 2 years of age and levels out around 28 points after 5 years of age. The unpaired Student’s t-test showed a statistically significant difference between neurotypical and ASD children at each age bin (p<0.0001).

**Figure 1.**
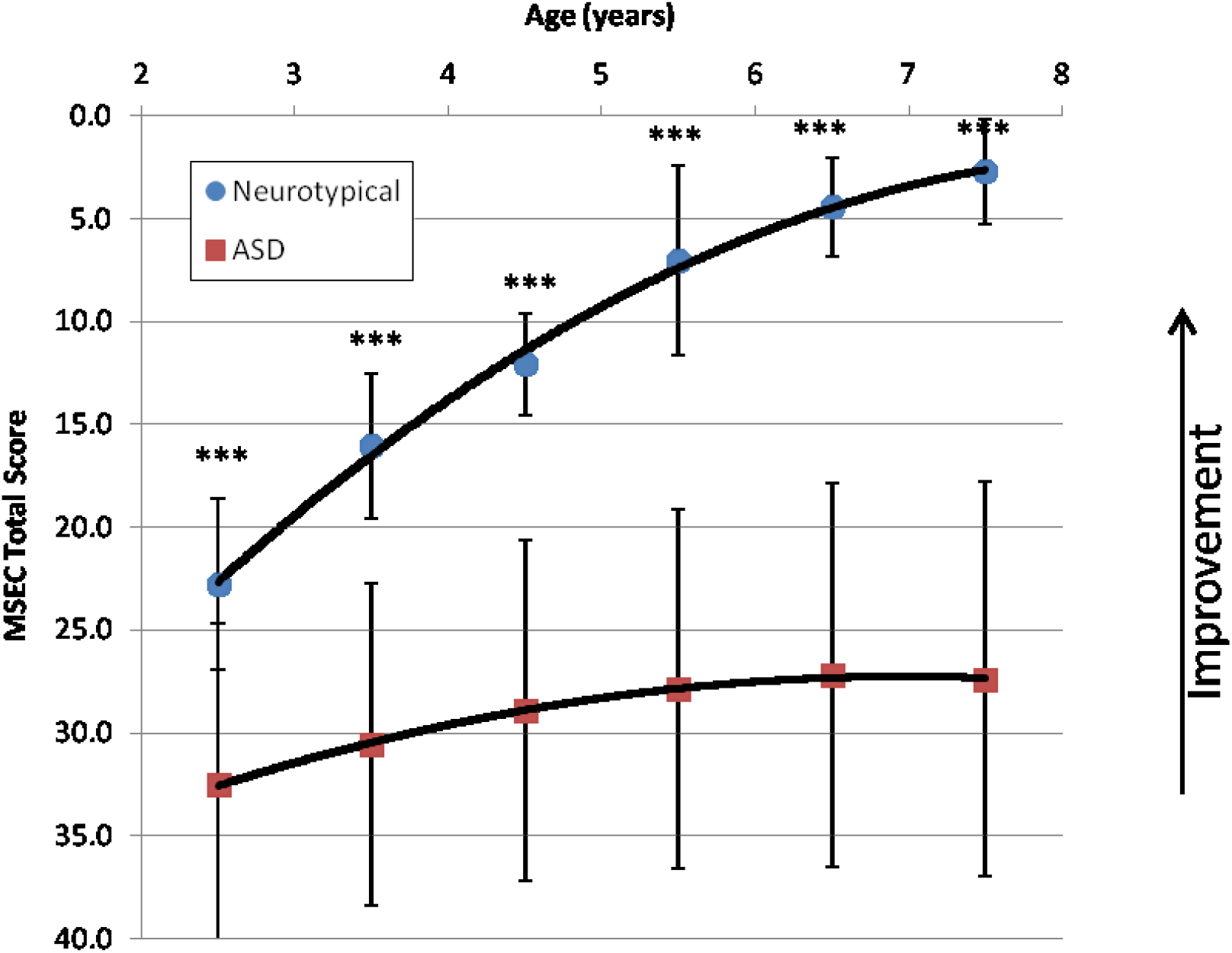
Scatter plot of the total MSEC score as a function of age in ASD children (squares) and neurotypical children (circles). P-value is marked: ***<0.0001.

To test the hypothesis that MSEC could be a screening test used for the early diagnosis of PFS problems, we have constructed receiver operating characteristic curves for each age group, Figure 2. Using a MSEC score cutoff of 29 at 2 years of age separated ASD from neurotypical children with a sensitivity 0.77 and a specificity 0.88 (Table S1). Furthermore, the differences between the groups increases with age. Using a MSEC score cutoff of 21 at 3 years of age separated ASD from neurotypical children with a sensitivity of 0.87 and a specificity of 0.95 (Table S2); at 4 years of age, a MSEC score cutoff of 17 separated the groups with a sensitivity of 0.93 and a specificity of 1.0 (Table S3); at 5 years of age, a MSEC score cutoff of 16 separated the groups with a sensitivity of 0.91 and a specificity of 0.96 (Table S4); at 6 years of age, a MSEC score cutoff of 11 separated the groups with a sensitivity of 0.94 and a specificity of 1.0 (Table S5); at 7 years of age, a MSEC score cutoff of 10 separated the groups with a sensitivity of 0.95 and a specificity of 1.0 (Table S6).

**Figure 2.**
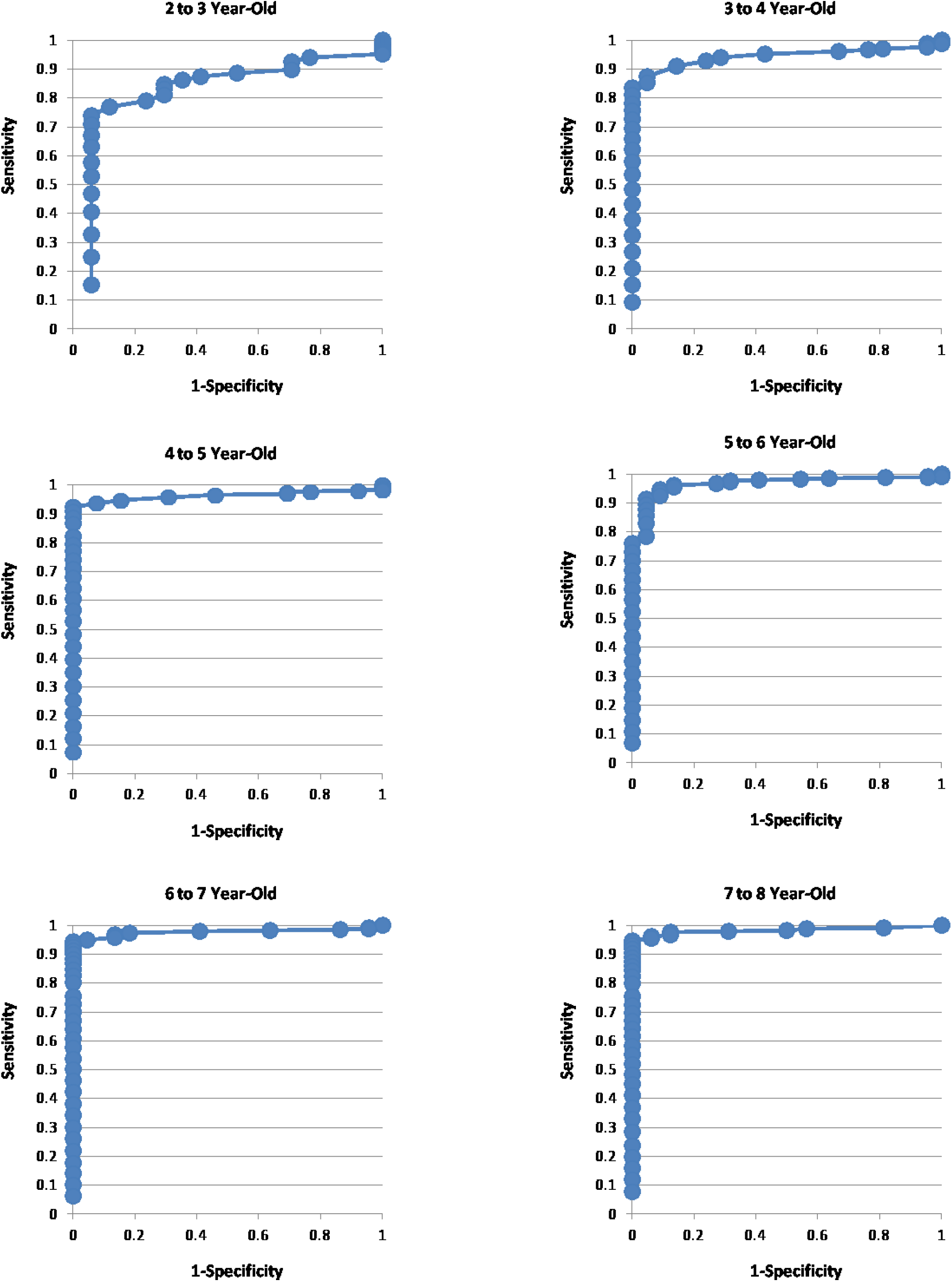
Receiver Operating Characteristic (ROC) curves for MSEC score.

Individual MSEC items provide details on developmental differences between neurotypical and ASD children. Table 3 shows the percentage of parents answering “very true” in each MSEC item and Table 3 shows the same information for children with ASD. All neurotypical children over 3 years of age but only 15% of ASD children understood simple stories that were read aloud (increasing to the maximum of 21% after 5 years of age; Table 3 and 4, item 1). 85% of neurotypical children 4 years of age understood elaborate fairy tales that were read aloud (item 2) as compared to only 9% of ASD children of the same age (increasing to the maximum of 11% after 5 years of age). 81% of neurotypical children 4 years of age understood explanations about people, objects or situations beyond the immediate surroundings (item 20) as compared to only 8% of ASD children of the same age (increasing to the maximum of 14% after 6 years of age).

**Table 3.**
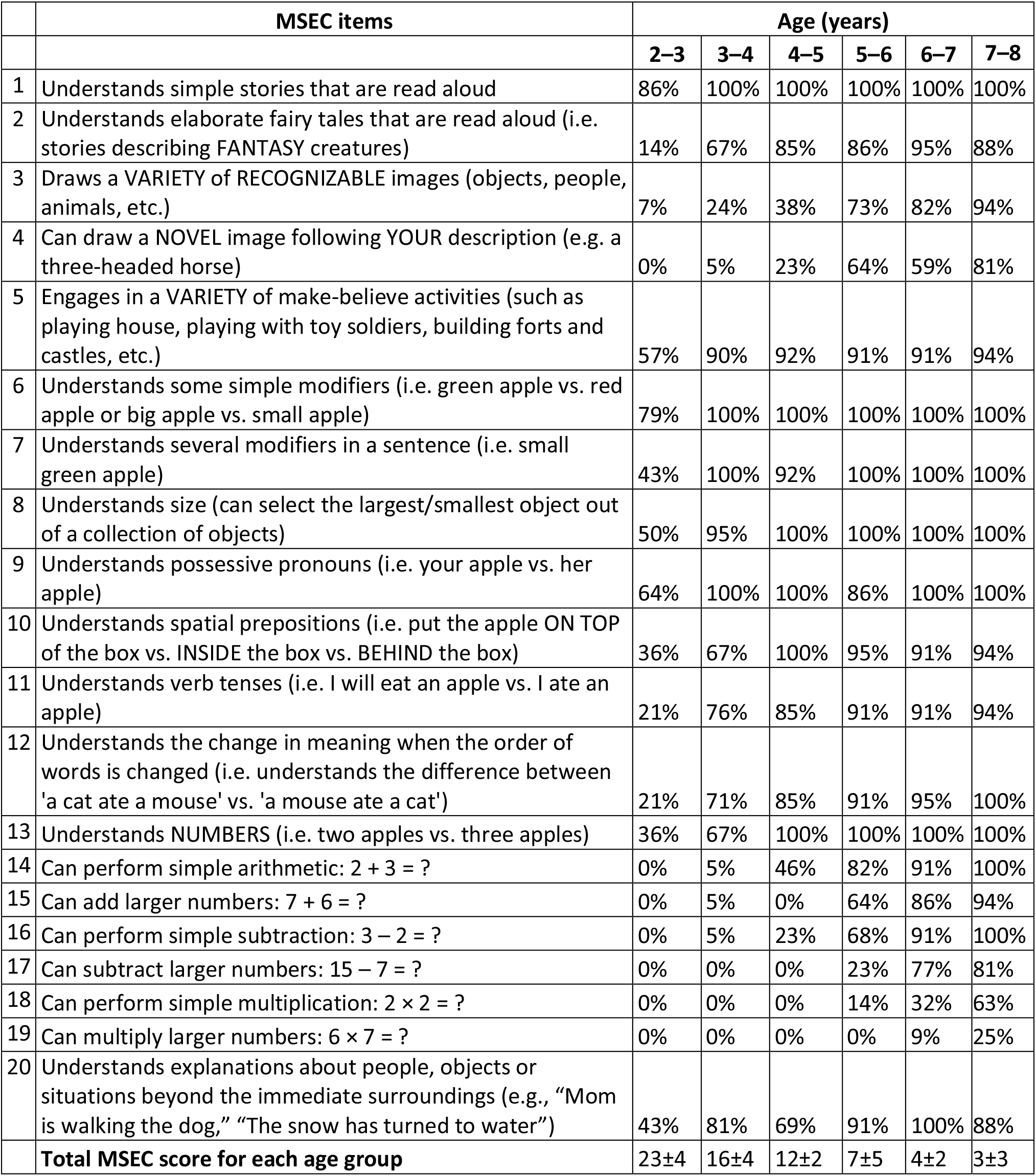
Percentage of parents answering “very true” for each MSEC item in neurotypical children.

|  | MSEC items | Age (years) |  |  |  |  |  |
| --- | --- | --- | --- | --- | --- | --- | --- |
|  |  | 2–3 | 3–4 | 4–5 | 5–6 | 6–7 | 7–8 |
| 1 | Understands simple stories that are read aloud | 86% | 100% | 100% | 100% | 100% | 100% |
| 2 | Understands elaborate fairy tales that are read aloud (i.e. stories describing FANTASY creatures) | 14% | 67% | 85% | 86% | 95% | 88% |
| 3 | Draws a VARIETY of RECOGNIZABLE images (objects, people, animals, etc.) | 7% | 24% | 38% | 73% | 82% | 94% |
| 4 | Can draw a NOVEL image following YOUR description (e.g. a three-headed horse) | 0% | 5% | 23% | 64% | 59% | 81% |
| 5 | Engages in a VARIETY of make-believe activities (such as playing house, playing with toy soldiers, building forts and castles, etc.) | 57% | 90% | 92% | 91% | 91% | 94% |
| 6 | Understands some simple modifiers (i.e. green apple vs. red apple or big apple vs. small apple) | 79% | 100% | 100% | 100% | 100% | 100% |
| 7 | Understands several modifiers in a sentence (i.e. small green apple) | 43% | 100% | 92% | 100% | 100% | 100% |
| 8 | Understands size (can select the largest/smallest object out of a collection of objects) | 50% | 95% | 100% | 100% | 100% | 100% |
| 9 | Understands possessive pronouns (i.e. your apple vs. her apple) | 64% | 100% | 100% | 86% | 100% | 100% |
| 10 | Understands spatial prepositions (i.e. put the apple ON TOP of the box vs. INSIDE the box vs. BEHIND the box) | 36% | 67% | 100% | 95% | 91% | 94% |
| 11 | Understands verb tenses (i.e. I will eat an apple vs. I ate an apple) | 21% | 76% | 85% | 91% | 91% | 94% |
| 12 | Understands the change in meaning when the order of words is changed (i.e. understands the difference between 'a cat ate a mouse' vs. 'a mouse ate a cat') | 21% | 71% | 85% | 91% | 95% | 100% |
| 13 | Understands NUMBERS (i.e. two apples vs. three apples) | 36% | 67% | 100% | 100% | 100% | 100% |
| 14 | Can perform simple arithmetic: $2 + 3 = ?$ | 0% | 5% | 46% | 82% | 91% | 100% |
| 15 | Can add larger numbers: $7 + 6 = ?$ | 0% | 5% | 0% | 64% | 86% | 94% |
| 16 | Can perform simple subtraction: $3 - 2 = ?$ | 0% | 5% | 23% | 68% | 91% | 100% |
| 17 | Can subtract larger numbers: $15 - 7 = ?$ | 0% | 0% | 0% | 23% | 77% | 81% |
| 18 | Can perform simple multiplication: $2 \times 2 = ?$ | 0% | 0% | 0% | 14% | 32% | 63% |
| 19 | Can multiply larger numbers: $6 \times 7 = ?$ | 0% | 0% | 0% | 0% | 9% | 25% |
| 20 | Understands explanations about people, objects or situations beyond the immediate surroundings (e.g., “Mom is walking the dog,” “The snow has turned to water”) | 43% | 81% | 69% | 91% | 100% | 88% |
|  | **Total MSEC score for each age group** | 23±4 | 16±4 | 12±2 | 7±5 | 4±2 | 3±3 |

**Table 4.**
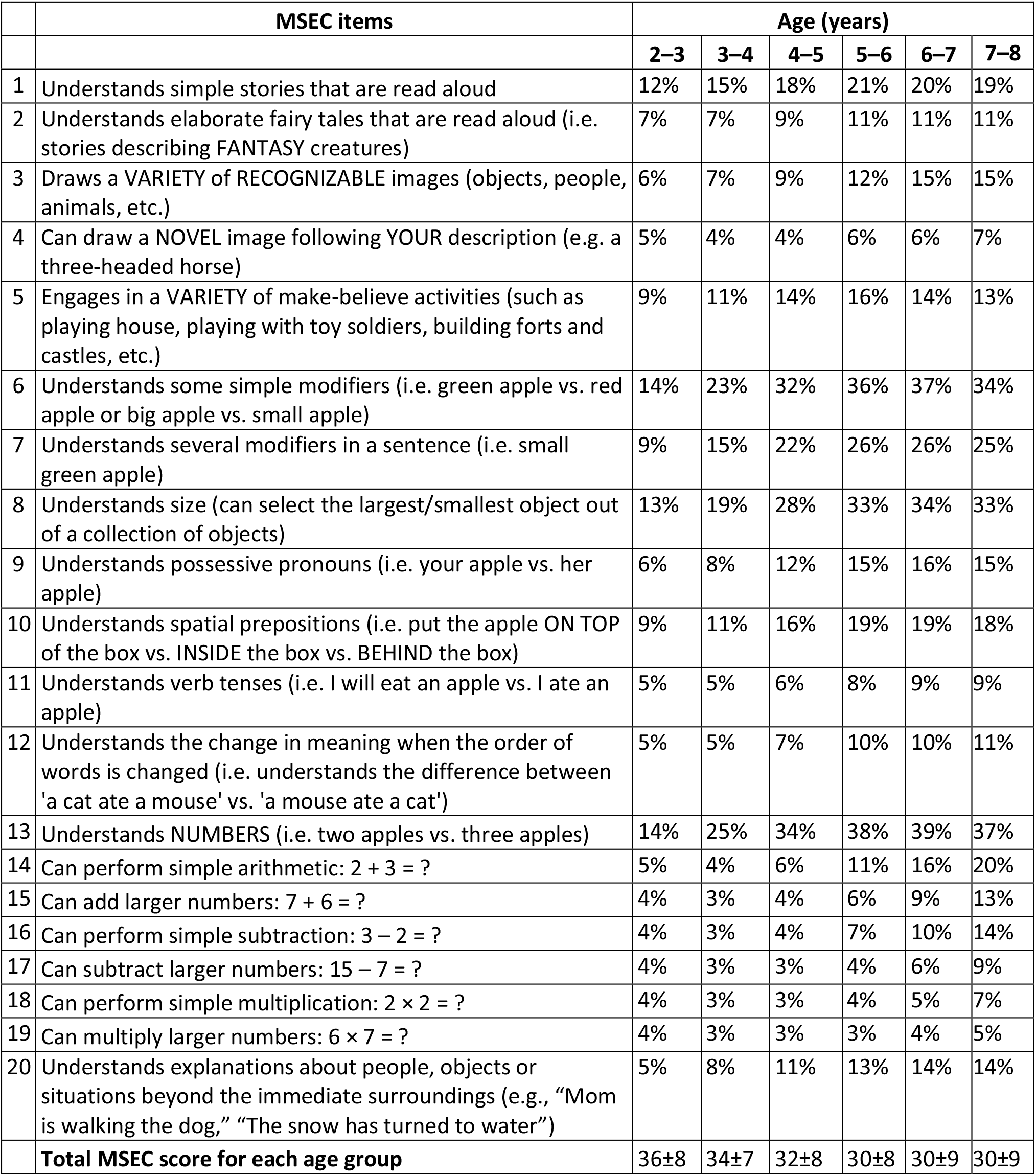
Percentage of parents answering “very true” for each MSEC item in children with ASD.

|  | MSEC items | Age (years) |  |  |  |  |  |
| --- | --- | --- | --- | --- | --- | --- | --- |
|  |  | 2–3 | 3–4 | 4–5 | 5–6 | 6–7 | 7–8 |
| 1 | Understands simple stories that are read aloud | 12% | 15% | 18% | 21% | 20% | 19% |
| 2 | Understands elaborate fairy tales that are read aloud (i.e. stories describing FANTASY creatures) | 7% | 7% | 9% | 11% | 11% | 11% |
| 3 | Draws a VARIETY of RECOGNIZABLE images (objects, people, animals, etc.) | 6% | 7% | 9% | 12% | 15% | 15% |
| 4 | Can draw a NOVEL image following YOUR description (e.g. a three-headed horse) | 5% | 4% | 4% | 6% | 6% | 7% |
| 5 | Engages in a VARIETY of make-believe activities (such as playing house, playing with toy soldiers, building forts and castles, etc.) | 9% | 11% | 14% | 16% | 14% | 13% |
| 6 | Understands some simple modifiers (i.e. green apple vs. red apple or big apple vs. small apple) | 14% | 23% | 32% | 36% | 37% | 34% |
| 7 | Understands several modifiers in a sentence (i.e. small green apple) | 9% | 15% | 22% | 26% | 26% | 25% |
| 8 | Understands size (can select the largest/smallest object out of a collection of objects) | 13% | 19% | 28% | 33% | 34% | 33% |
| 9 | Understands possessive pronouns (i.e. your apple vs. her apple) | 6% | 8% | 12% | 15% | 16% | 15% |
| 10 | Understands spatial prepositions (i.e. put the apple ON TOP of the box vs. INSIDE the box vs. BEHIND the box) | 9% | 11% | 16% | 19% | 19% | 18% |
| 11 | Understands verb tenses (i.e. I will eat an apple vs. I ate an apple) | 5% | 5% | 6% | 8% | 9% | 9% |
| 12 | Understands the change in meaning when the order of words is changed (i.e. understands the difference between 'a cat ate a mouse' vs. 'a mouse ate a cat') | 5% | 5% | 7% | 10% | 10% | 11% |
| 13 | Understands NUMBERS (i.e. two apples vs. three apples) | 14% | 25% | 34% | 38% | 39% | 37% |
| 14 | Can perform simple arithmetic: $2 + 3 = ?$ | 5% | 4% | 6% | 11% | 16% | 20% |
| 15 | Can add larger numbers: $7 + 6 = ?$ | 4% | 3% | 4% | 6% | 9% | 13% |
| 16 | Can perform simple subtraction: $3 - 2 = ?$ | 4% | 3% | 4% | 7% | 10% | 14% |
| 17 | Can subtract larger numbers: $15 - 7 = ?$ | 4% | 3% | 3% | 4% | 6% | 9% |
| 18 | Can perform simple multiplication: $2 \times 2 = ?$ | 4% | 3% | 3% | 4% | 5% | 7% |
| 19 | Can multiply larger numbers: $6 \times 7 = ?$ | 4% | 3% | 3% | 3% | 4% | 5% |
| 20 | Understands explanations about people, objects or situations beyond the immediate surroundings (e.g., “Mom is walking the dog,” “The snow has turned to water”) | 5% | 8% | 11% | 13% | 14% | 14% |
|  | **Total MSEC score for each age group** | 36±8 | 34±7 | 32±8 | 30±8 | 30±9 | 30±9 |

Understanding of noun-adjective combinations, spatial prepositions, and flexible syntax was similarly different between neurotypical and ASD children. Simple modifiers (item 6) were understood by all neurotypical children at 3 years of age, but only by 23% of ASD children at the same age (increasing to the maximum of 37% after 6 years of age). Using several modifiers together in a sentence (item 7) was also understood by all neurotypical children at 3 years of age, but only by 15% of ASD children of the same age (increasing to the maximum of 26% after 5 years of age). Superlatives (item 8) were understood by nearly all neurotypical children at 3 years of age, but only by 19% of ASD children of the same age (increasing to the maximum of 34% after 6 years of age). Possessive pronouns (item 9) were understood by all neurotypical children at 3 years of age, but only by 8% of ASD children of the same age (increasing to the maximum of 16% after 6 years of age). Spatial prepositions (item 10) were understood by all neurotypical children at 4 years of age, but only by 16% of ASD children of the same age (increasing to the maximum of 19% after 5 years of age). Verb tenses (item 11) were understood by 85% of neurotypical children at 4 years of age, but only by 6% of ASD children of the same age (increasing to the maximum of 9% after 9 years of age). Syntax (item 12) was understood by 85% of neurotypical children at 4 years of age, but only by 7% of ASD children of the same age (increasing to the maximum of 11% after 7 years of age).

Drawing a variety of recognizable images (item 3) was reported in 94% of neurotypical children at 7 years of age, compared to 15% of ASD children of the same age. The ability to draw a novel image following a complex description (e.g., a three-headed horse; item 4) was reported in 81% of neurotypical children at 7 years of age, compared to 7% of ASD children of the same age. Furthermore, 90% of neurotypical children engaged in a variety of make-believe activities (item 5) at 3 years of age, compared to only 11% of ASD children at the same age (increasing to the maximum of 16% after 5 years of age).

Most neurotypical children acquired first arithmetic skills (item 14) around 4 years of age, were able to perform simple subtraction (item 16) around 5 years of age, and were able to perform simple multiplication (item 18) around 6 years of age. Among ASD children 7 years of age only 21% were able to perform simple addition, 15% simple subtraction, and 8% simple multiplication.

## Discussion

This manuscript compares combinatorial language acquisition between 111 neurotypical individuals, whose parents completed an online Google form and 29138 ASD children, whose parents completed the MSEC evaluation within a language training app. MSEC revealed significant differences between normally developing and ASD children as early as 2 years of age (p<0.0001) (Figure 1). The trajectory of MSEC score was also different between the groups. In neurotypical children the MSEC total score improves linearly with age, from 22.8±4.2 points at 2 years of age to 2.7±2.5 points at 7 years of age. Conversely, In ASD children the MSEC total score starts at 32.5±7.9 points at 2 years of age and levels out around 28 points after 5 years of age.

The MSEC score cutoff of 29 separates ASD from neurotypical children with high sensitivity (0.77) and high specificity (0.88) at 2 years of age (Figure 2). As expected, the difference between the groups increases with age: a MSEC score cutoff of 21 at 3 years of age separated ASD from neurotypical children with a sensitivity of 0.87 and a specificity of 0.95; at 4 years of age, a MSEC score cutoff of 17 separated the groups with a sensitivity of 0.93 and a specificity of 1.0; at 5 years of age, a MSEC score cutoff of 16 separated the groups with a sensitivity of 0.91 and a specificity of 0.96; at 6 years of age, a MSEC score cutoff of 11 separated the groups with a sensitivity of 0.94 and a specificity of 1.0; at 7 years of age, a MSEC score cutoff of 10 separated the groups with a sensitivity of 0.95 and a specificity of 1.0. We conclude that MSEC can be used to detect combinatorial language problems as early as 2 years of age.

Neurotypical developmental trajectories revealed by individual MSEC items fit well into literature on normal child development (Table 3). Congruent with previous studies, a majority of neurotypical children were reported to display pretend play, understand simple stories, possessive pronouns, and simple modifiers before 3 years of age, to understand elaborate fairy tales, explanations about people, several modifiers in a sentence, size, spatial prepositions, verb tenses, syntax, and numbers by 4 years of age, to grasp the concept of addition by 5 years of age, subtraction by 6 years of age, and multiplication by 7 years of age (Piaget, 1965; Prather & Alibali, 2011; Rice et al., 1998; Rieber & Carton, 1987; Washington & Naremore, 1978). ASD children exhibit a clear delay in every MSEC item (Table 4).

A threshold indicating good PFS ability can be identified in the MSEC scale similar to the previously determined threshold 7 out of 10 on the Linguistic Evaluation of Prefrontal Synthesis (LEPS), the clinician-assessed test that measures the same concept as the parent-report MSEC (Vyshedskiy, DuBois, et al., 2020). The LEPS score of 7/10 and the MSEC score of 12/40 are reached by typically developing children at 4 years of age. Thus, a LEPS score of ≥7/10 and an MSEC score of ≤12/40 signify acquisition of the PFS ability. Based on results presented in this manuscript among neurotypical children 5 years and older, 97% reached the threshold for good PFS ability (MSEC score 12 or lower). Surprisingly, only 6% of ASD children reached the threshold for good PFS ability at any age, indicating the scale of the PFS paralysis problem and underscoring the importance of regular assessments of PFS and combinatorial language.

### Longitudinal intervention studies

Longitudinal intervention studies commonly use parent-reported assessments of child development as an outcome measure (Ebert, 2017; Sikich et al., 2021). Parent-report tools for tracking expressive language, sociability, physical development, and health have been available for a long time (Rimland & Edelson, 1999), however, parent-report tools for monitoring combinatorial receptive language have been absent. Since combinatorial receptive language acquisition in atypically developing children can be dissociated from expressive language, it is essential to monitor combinatorial receptive language separately from expressive language.

In two recent studies, the MSEC demonstrated an ability to measure an effect different from expressive language. In a 3-year clinical study of 6454 children with ASD, children who engaged with a novel language intervention increased their MSEC score 2.2-fold when compared to children with similar initial evaluations (*p*<0.0001) (Vyshedskiy, DuBois, et al., 2020). At the same time, the difference between the groups in the expressive language score was only 1.4-fold (*p*=0.0144). In another 3-year epidemiological study of the effect of passive video and television watching on children with ASD (N= 3227), longer video and television watching were associated with slightly better development of expressive language but significantly impeded development of receptive language, as measured by the MSEC (Fridberg et al., 2021). This dissociation between receptive and expressive language trajectories observed in both studies strongly imply the need for a regular assessment of combinatorial receptive language in addition to expressive language. If the MSEC evaluation data were not available in the television study, we would have concluded that longer video and television watching time had a positive effect on children with ASD, as exhibited by the expressive language subscale (other subscales [sociability, sensory awareness, and health] showed no difference between the groups). Only with the addition of the MSEC scale were we able to detect the significant negative effect of passive video and television watching.

In addition to its usefulness in epidemiological studies, a combinatorial receptive language assessment, such as the MSEC, could aid parents in quantitative monitoring of their child’s language trajectory. Parent-report tools like ATEC and MSEC democratize child neurological assessment and empower parents to quantitatively monitor their children’s improvement over time.

Both MSEC and LEPS are copyright-free and can be used on their own or in combination with the four subscales of the ATEC evaluation (expressive language, sociability, sensory awareness, and health) (Mahapatra et al., 2018; Rimland & Edelson, 1999). As both, MSEC and LEPS, do not rely on productive language, they can also be used in nonverbal and minimally verbal children.

## Data Availability

All data produced in the present study are available upon reasonable request to the authors

## Author contributions

AV designed the study. AV and MA analyzed the data. AV and MA wrote the paper.

## Informed Consent

Caregivers have consented to anonymized data analysis and publication of the results. The study was conducted in compliance with the Declaration of Helsinki ^39^.

## Compliance with Ethical Standards

Using the Department of Health and Human Services regulations found at 45 CFR 46.101(b)(4), the Biomedical Research Alliance of New York LLC Institutional Review Board (IRB) determined that this research project is exempt from IRB oversight.

## Data Availability

De-identified raw data from this manuscript are available from the corresponding author upon reasonable request.

## Code availability statement

Code is available from the corresponding author upon reasonable request.

