## Supplementary Material for "Combinatorial language parent-report score differs significantly between typically developing children and those with Autism Spectrum Disorders"

Table S1. Receiver operating characteristic curves for 2 to 3 year-old children. Using a MSEC score cutoff of 29 separated ASD from neurotypical children with a sensitivity 0.77 and a specificity 0.88.

| MSEC cutoff | Sensitivity | Specificity | 1-Specificity |
|---|---|---|---|
| 0 | 1.00 | 0.00 | 1.00 |
| 1 | 0.99 | 0.00 | 1.00 |
| 2 | 0.99 | 0.00 | 1.00 |
| 3 | 0.99 | 0.00 | 1.00 |
| 4 | 0.99 | 0.00 | 1.00 |
| 5 | 0.99 | 0.00 | 1.00 |
| 6 | 0.99 | 0.00 | 1.00 |
| 7 | 0.99 | 0.00 | 1.00 |
| 8 | 0.98 | 0.00 | 1.00 |
| 9 | 0.98 | 0.00 | 1.00 |
| 10 | 0.98 | 0.00 | 1.00 |
| 11 | 0.98 | 0.00 | 1.00 |
| 12 | 0.98 | 0.00 | 1.00 |
| 13 | 0.97 | 0.00 | 1.00 |
| 14 | 0.97 | 0.00 | 1.00 |
| 15 | 0.97 | 0.00 | 1.00 |
| 16 | 0.97 | 0.00 | 1.00 |
| 17 | 0.96 | 0.00 | 1.00 |
| 18 | 0.95 | 0.00 | 1.00 |
| 19 | 0.94 | 0.24 | 0.76 |
| 20 | 0.93 | 0.29 | 0.71 |
| 21 | 0.90 | 0.29 | 0.71 |
| 22 | 0.89 | 0.47 | 0.53 |
| 23 | 0.88 | 0.59 | 0.41 |
| 24 | 0.86 | 0.65 | 0.35 |
| 25 | 0.85 | 0.71 | 0.29 |
| 26 | 0.83 | 0.71 | 0.29 |
| 27 | 0.81 | 0.71 | 0.29 |
| 28 | 0.79 | 0.76 | 0.24 |
| **29** | **0.77** | **0.88** | **0.12** |
| 30 | 0.74 | 0.94 | 0.06 |
| 31 | 0.71 | 0.94 | 0.06 |
| 32 | 0.67 | 0.94 | 0.06 |
| 33 | 0.63 | 0.94 | 0.06 |
| 34 | 0.58 | 0.94 | 0.06 |
| 35 | 0.53 | 0.94 | 0.06 |
| 36 | 0.47 | 0.94 | 0.06 |
| 37 | 0.40 | 0.94 | 0.06 |
| 38 | 0.33 | 0.94 | 0.06 |
| 39 | 0.25 | 0.94 | 0.06 |
| 40 | 0.15 | 0.94 | 0.06 |

Table S2. Receiver operating characteristic curves for 3 to 4 year-old children. Using a MSEC score cutoff of 21 separated ASD from neurotypical children with a sensitivity 0.87 and a specificity 0.95.

| MSEC cutoff | Sensitivity | Specificity | 1-Specificity |
|---|---|---|---|
| 0 | 1.00 | 0.00 | 1.00 |
| 1 | 0.99 | 0.00 | 1.00 |
| 2 | 0.99 | 0.00 | 1.00 |
| 3 | 0.99 | 0.00 | 1.00 |
| 4 | 0.99 | 0.00 | 1.00 |
| 5 | 0.99 | 0.00 | 1.00 |
| 6 | 0.99 | 0.05 | 0.95 |
| 7 | 0.99 | 0.05 | 0.95 |
| 8 | 0.99 | 0.05 | 0.95 |
| 9 | 0.99 | 0.05 | 0.95 |
| 10 | 0.98 | 0.05 | 0.95 |
| 11 | 0.98 | 0.05 | 0.95 |
| 12 | 0.98 | 0.05 | 0.95 |
| 13 | 0.98 | 0.05 | 0.95 |
| 14 | 0.97 | 0.19 | 0.81 |
| 15 | 0.97 | 0.24 | 0.76 |
| 16 | 0.96 | 0.33 | 0.67 |
| 17 | 0.95 | 0.57 | 0.43 |
| 18 | 0.94 | 0.71 | 0.29 |
| 19 | 0.93 | 0.76 | 0.24 |
| 20 | 0.91 | 0.86 | 0.14 |
| **21** | 0.87 | 0.95 | 0.05 |
| 22 | 0.85 | 0.95 | 0.05 |
| 23 | 0.84 | 1.00 | 0.00 |
| 24 | 0.81 | 1.00 | 0.00 |
| 25 | 0.78 | 1.00 | 0.00 |
| 26 | 0.76 | 1.00 | 0.00 |
| 27 | 0.73 | 1.00 | 0.00 |
| 28 | 0.69 | 1.00 | 0.00 |
| 29 | 0.66 | 1.00 | 0.00 |
| 30 | 0.62 | 1.00 | 0.00 |
| 31 | 0.58 | 1.00 | 0.00 |
| 32 | 0.54 | 1.00 | 0.00 |
| 33 | 0.49 | 1.00 | 0.00 |
| 34 | 0.43 | 1.00 | 0.00 |
| 35 | 0.38 | 1.00 | 0.00 |
| 36 | 0.33 | 1.00 | 0.00 |
| 37 | 0.27 | 1.00 | 0.00 |
| 38 | 0.21 | 1.00 | 0.00 |
| 39 | 0.15 | 1.00 | 0.00 |
| 40 | 0.09 | 1.00 | 0.00 |

Table S3. Receiver operating characteristic curves for 4 to 5 year-old children. Using a MSEC score cutoff of 17 separated ASD from neurotypical children with a sensitivity 0.93 and a specificity 1.0.

| MSEC cutoff | Sensitivity | Specificity | 1-Specificity |
|---|---|---|---|
| 0 | 1.00 | 0.00 | 1.00 |
| 1 | 0.99 | 0.00 | 1.00 |
| 2 | 0.99 | 0.00 | 1.00 |
| 3 | 0.99 | 0.00 | 1.00 |
| 4 | 0.99 | 0.00 | 1.00 |
| 5 | 0.99 | 0.00 | 1.00 |
| 6 | 0.99 | 0.00 | 1.00 |
| 7 | 0.99 | 0.00 | 1.00 |
| 8 | 0.98 | 0.00 | 1.00 |
| 9 | 0.98 | 0.08 | 0.92 |
| 10 | 0.98 | 0.23 | 0.77 |
| 11 | 0.97 | 0.31 | 0.69 |
| 12 | 0.97 | 0.31 | 0.69 |
| 13 | 0.97 | 0.54 | 0.46 |
| 14 | 0.96 | 0.69 | 0.31 |
| 15 | 0.95 | 0.85 | 0.15 |
| 16 | 0.94 | 0.92 | 0.08 |
| **17** | **0.93** | **1.00** | **0.00** |
| 18 | 0.91 | 1.00 | 0.00 |
| 19 | 0.89 | 1.00 | 0.00 |
| 20 | 0.87 | 1.00 | 0.00 |
| 21 | 0.82 | 1.00 | 0.00 |
| 22 | 0.80 | 1.00 | 0.00 |
| 23 | 0.77 | 1.00 | 0.00 |
| 24 | 0.74 | 1.00 | 0.00 |
| 25 | 0.71 | 1.00 | 0.00 |
| 26 | 0.68 | 1.00 | 0.00 |
| 27 | 0.64 | 1.00 | 0.00 |
| 28 | 0.61 | 1.00 | 0.00 |
| 29 | 0.57 | 1.00 | 0.00 |
| 30 | 0.53 | 1.00 | 0.00 |
| 31 | 0.48 | 1.00 | 0.00 |
| 32 | 0.44 | 1.00 | 0.00 |
| 33 | 0.40 | 1.00 | 0.00 |
| 34 | 0.35 | 1.00 | 0.00 |
| 35 | 0.30 | 1.00 | 0.00 |
| 36 | 0.26 | 1.00 | 0.00 |
| 37 | 0.21 | 1.00 | 0.00 |
| 38 | 0.17 | 1.00 | 0.00 |
| 39 | 0.12 | 1.00 | 0.00 |
| 40 | 0.07 | 1.00 | 0.00 |

Table S4. Receiver operating characteristic curves for 5 to 6 year-old children. Using a MSEC score cutoff of 16 separated ASD from neurotypical children with a sensitivity 0.91 and a specificity 0.96.

| MSEC cutoff | Sensitivity | Specificity | 1-Specificity |
|---|---|---|---|
| 0 | 1.00 | 0.00 | 1.00 |
| 1 | 0.99 | 0.00 | 1.00 |
| 2 | 0.99 | 0.05 | 0.95 |
| 3 | 0.99 | 0.05 | 0.95 |
| 4 | 0.99 | 0.18 | 0.82 |
| 5 | 0.99 | 0.36 | 0.64 |
| 6 | 0.98 | 0.45 | 0.55 |
| 7 | 0.98 | 0.59 | 0.41 |
| 8 | 0.98 | 0.68 | 0.32 |
| 9 | 0.97 | 0.68 | 0.32 |
| 10 | 0.97 | 0.73 | 0.27 |
| 11 | 0.96 | 0.86 | 0.14 |
| 12 | 0.96 | 0.86 | 0.14 |
| 13 | 0.95 | 0.91 | 0.09 |
| 14 | 0.94 | 0.91 | 0.09 |
| 15 | 0.93 | 0.91 | 0.09 |
| **16** | **0.91** | **0.95** | **0.05** |
| 17 | 0.90 | 0.95 | 0.05 |
| 18 | 0.88 | 0.95 | 0.05 |
| 19 | 0.86 | 0.95 | 0.05 |
| 20 | 0.83 | 0.95 | 0.05 |
| 21 | 0.78 | 0.95 | 0.05 |
| 22 | 0.76 | 1.00 | 0.00 |
| 23 | 0.73 | 1.00 | 0.00 |
| 24 | 0.70 | 1.00 | 0.00 |
| 25 | 0.67 | 1.00 | 0.00 |
| 26 | 0.63 | 1.00 | 0.00 |
| 27 | 0.60 | 1.00 | 0.00 |
| 28 | 0.56 | 1.00 | 0.00 |
| 29 | 0.52 | 1.00 | 0.00 |
| 30 | 0.48 | 1.00 | 0.00 |
| 31 | 0.44 | 1.00 | 0.00 |
| 32 | 0.39 | 1.00 | 0.00 |
| 33 | 0.35 | 1.00 | 0.00 |
| 34 | 0.31 | 1.00 | 0.00 |
| 35 | 0.26 | 1.00 | 0.00 |
| 36 | 0.23 | 1.00 | 0.00 |
| 37 | 0.19 | 1.00 | 0.00 |
| 38 | 0.15 | 1.00 | 0.00 |
| 39 | 0.11 | 1.00 | 0.00 |
| 40 | 0.07 | 1.00 | 0.00 |

Table S5. Receiver operating characteristic curves for 6 to 7 year-old children. Using a MSEC score cutoff of 11 separated ASD from neurotypical children with a sensitivity 0.94 and a specificity 1.0.

| MSEC cutoff | Sensitivity | Specificity | 1-Specificity |
|---|---|---|---|
| 0 | 1.00 | 0.00 | 1.00 |
| 1 | 0.99 | 0.05 | 0.95 |
| 2 | 0.99 | 0.05 | 0.95 |
| 3 | 0.98 | 0.14 | 0.86 |
| 4 | 0.98 | 0.36 | 0.64 |
| 5 | 0.98 | 0.59 | 0.41 |
| 6 | 0.97 | 0.82 | 0.18 |
| 7 | 0.97 | 0.86 | 0.14 |
| 8 | 0.96 | 0.86 | 0.14 |
| 9 | 0.96 | 0.86 | 0.14 |
| 10 | 0.95 | 0.95 | 0.05 |
| **11** | **0.94** | **1.00** | **0.00** |
| 12 | 0.93 | 1.00 | 0.00 |
| 13 | 0.92 | 1.00 | 0.00 |
| 14 | 0.92 | 1.00 | 0.00 |
| 15 | 0.90 | 1.00 | 0.00 |
| 16 | 0.88 | 1.00 | 0.00 |
| 17 | 0.87 | 1.00 | 0.00 |
| 18 | 0.85 | 1.00 | 0.00 |
| 19 | 0.83 | 1.00 | 0.00 |
| 20 | 0.80 | 1.00 | 0.00 |
| 21 | 0.75 | 1.00 | 0.00 |
| 22 | 0.73 | 1.00 | 0.00 |
| 23 | 0.70 | 1.00 | 0.00 |
| 24 | 0.67 | 1.00 | 0.00 |
| 25 | 0.64 | 1.00 | 0.00 |
| 26 | 0.61 | 1.00 | 0.00 |
| 27 | 0.58 | 1.00 | 0.00 |
| 28 | 0.54 | 1.00 | 0.00 |
| 29 | 0.50 | 1.00 | 0.00 |
| 30 | 0.46 | 1.00 | 0.00 |
| 31 | 0.42 | 1.00 | 0.00 |
| 32 | 0.38 | 1.00 | 0.00 |
| 33 | 0.34 | 1.00 | 0.00 |
| 34 | 0.30 | 1.00 | 0.00 |
| 35 | 0.26 | 1.00 | 0.00 |
| 36 | 0.22 | 1.00 | 0.00 |
| 37 | 0.18 | 1.00 | 0.00 |
| 38 | 0.14 | 1.00 | 0.00 |
| 39 | 0.10 | 1.00 | 0.00 |
| 40 | 0.06 | 1.00 | 0.00 |

Table S6. Receiver operating characteristic curves for 7 to 8 year-old children. Using a MSEC score cutoff of 10 separated ASD from neurotypical children with a sensitivity 0.95 and a specificity 1.0.

| MSEC cutoff | Sensitivity | Specificity | 1-Specificity |
|---|---|---|---|
| 0 | 1.00 | 0.00 | 1.00 |
| 1 | 0.99 | 0.19 | 0.81 |
| 2 | 0.99 | 0.44 | 0.56 |
| 3 | 0.98 | 0.50 | 0.50 |
| 4 | 0.98 | 0.69 | 0.31 |
| 5 | 0.98 | 0.88 | 0.13 |
| 6 | 0.97 | 0.88 | 0.13 |
| 7 | 0.97 | 0.88 | 0.13 |
| 8 | 0.96 | 0.94 | 0.06 |
| 9 | 0.96 | 0.94 | 0.06 |
| **10** | **0.95** | **1.00** | **0.00** |
| 11 | 0.94 | 1.00 | 0.00 |
| 12 | 0.93 | 1.00 | 0.00 |
| 13 | 0.92 | 1.00 | 0.00 |
| 14 | 0.90 | 1.00 | 0.00 |
| 15 | 0.89 | 1.00 | 0.00 |
| 16 | 0.87 | 1.00 | 0.00 |
| 17 | 0.86 | 1.00 | 0.00 |
| 18 | 0.84 | 1.00 | 0.00 |
| 19 | 0.82 | 1.00 | 0.00 |
| 20 | 0.80 | 1.00 | 0.00 |
| 21 | 0.75 | 1.00 | 0.00 |
| 22 | 0.72 | 1.00 | 0.00 |
| 23 | 0.70 | 1.00 | 0.00 |
| 24 | 0.67 | 1.00 | 0.00 |
| 25 | 0.64 | 1.00 | 0.00 |
| 26 | 0.62 | 1.00 | 0.00 |
| 27 | 0.58 | 1.00 | 0.00 |
| 28 | 0.55 | 1.00 | 0.00 |
| 29 | 0.52 | 1.00 | 0.00 |
| 30 | 0.49 | 1.00 | 0.00 |
| 31 | 0.45 | 1.00 | 0.00 |
| 32 | 0.41 | 1.00 | 0.00 |
| 33 | 0.37 | 1.00 | 0.00 |
| 34 | 0.33 | 1.00 | 0.00 |
| 35 | 0.29 | 1.00 | 0.00 |
| 36 | 0.24 | 1.00 | 0.00 |
| 37 | 0.20 | 1.00 | 0.00 |
| 38 | 0.16 | 1.00 | 0.00 |
| 39 | 0.12 | 1.00 | 0.00 |
| 40 | 0.08 | 1.00 | 0.00 |
